# Age-structured SIR model and resource growth dynamics: A preliminary COVID-19 study

**DOI:** 10.1101/2020.09.07.20184887

**Authors:** S. G. Babajanyan, Kang Hao Cheong

**Affiliations:** Science, Mathematics & Technology Cluster, Singapore University of Technology and Design, 8 Somapah Road, Singapore 487372

## Abstract

In this paper, we discuss three different response strategies to a disease outbreak and their economic implications in an age-structured population. We have utilized the classical age structured SIR-model, thus assuming that recovered people will not be infected again. Available resource dynamics is governed by the well-known logistic growth model, in which the reproduction coefficient depends on the disease outbreak spreading dynamics. We further investigate the feedback interaction of the disease spread dynamics and resource growth dynamics with the premise that the quality of treatment depends on the current economic situation. The very inclusion of mortality rates and economic considerations in the same model may be incongruous under certain positions, but in this model, we take a ‘realpolitik’ approach by exploring all of these factors together as it is done in reality.

## Introduction

The novel coronavirus (COVID-19) disease outbreak has been widespread among different countries causing more than 25 million cases of infection and more than 800K deaths. Despite the ongoing efforts, the vaccine to the disease is currently not available yet. In the absence of the vaccine, different countries have imposed various mitigation measures to either prevent the spread of COVID-19 or slow down the spread of the virus until the vaccine arrives.

The proposed mitigation measures are based on reducing the physical activities of the population [1, 2, 3, 4, 5, 6, 7, 8, 9, 10]. The classical Susceptible-Infectious-Recovered (SIR) model [11, 12, 13, 14] and several extensions of it have been used for modeling the virus spread and for understanding the impacts of different mitigation measures on the virus spreading. The possible economic impacts caused by mitigation measures and the optimal strategies for dealing with the disease outbreak have been suggested in various contexts [15, 16, 17, 18, 19, 20, 21, 22, 23]. Here, we investigate the feedback interaction of disease transmission and resource availability in the worst-case scenario: unavailability of the vaccine. We assume that neither a vaccine nor effective antiviral drugs become available. We also discuss disease outbreak transmission in an age-structured population: young and old group. The latter group is assumed to be more vulnerable to the infection: the death rate is higher for this group [1].

We discuss on one side the economic effects on the resources caused by a disease outbreak and the response strategies to it, and on the other side the impact of economic situation on the consequences of disease outbreak. The former effect is straightforward since the response strategies will have a direct impact on wealth growth. The latter effect illustrates the quality of treatment inside the hospitals. We model this phenomenon by assuming wealth dependent death rates inside the hospitals, that is, depleting the resources will cause an increase in deaths.

We assume that, in the absence of disease outbreak, the wealth growth is given by the logistic growth model known from the population dynamics with constant reproduction coefficient and carrying capacity. The reproduction coefficient consists of two parts: wealth generation and consumption. Wealth generation is due to the intragroup interactions in the first group. During the disease outbreak, only the agents from the suspected and recovered subgroups from the young group of population are contributing to the wealth growth. The consumption part has a negative impact on the reproduction coefficient of wealth growth dynamics. We assume that the consumption rate is the same throughout the population. Furthermore, we assume that infected people inside the hospitals will lead to additional treatment costs drawn from public resources. The impact of the mitigation measures is incorporated via the reproduction coefficient of available resources.

In this paper, we discuss three different response strategies for the disease outbreak: total lockdown, partial lockdown, and aiming towards achieving herd immunity as part of the SIR epidemic model. These strategies differ by their goals and their implementation methods. We then present results for the evolving COVID-19 pandemic under our theoretical model and assumptions. In the next section, we introduce the age-structured SIRD (suspected, infected, recovered, and dead) compartment model, the resource growth dynamics, feedback mechanism, and the calibration of various parameters. We will then discuss the mitigation strategies and their results in the last section.

## Model

### SIR model and wealth dynamics

Consider two population of agents 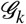, *k* = 1, 2. In each group there are subgroups *S_k_* (susceptible), *I_k_* (infected), *R_k_* (recovered) and *D_k_* (deaths), with transitions

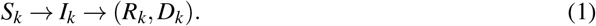

A single agent does not change its group (hence we denote this agent as 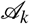), but it can change its subgroups. Denote by *p*(*S_k_*) the probability of 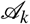 to be in *S_k_*. For each 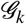 the probabilities obey the Markov equation:

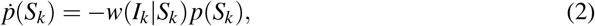

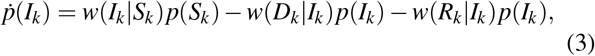

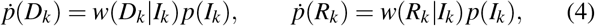

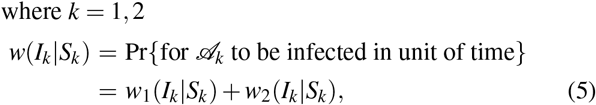

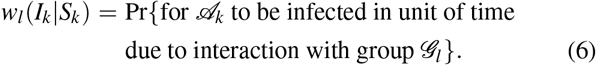

In addition to probabilities we introduce the number of agents *N(S_k_*) in subgroup *S_k_*, hence

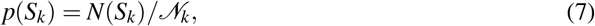

where 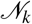 is the total number of agents in 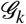. We stress that 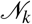 is conserved in time.

We also introduce

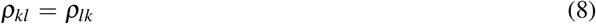

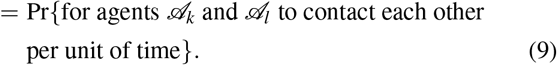

Hence we have in (5):

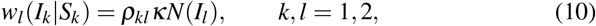

e.g. the probability for 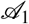 to get infected due to interaction with 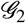 amounts to probability (per unit of time) *ρ*_12_ for contacting 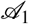 with 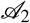, times the number of cases *N*(*I*_2_) that can lead to infection, *κ* is the probability of infection transmission.

Starting from (8) we can calculate the average number of contacts (per unit of time) between agents from within subgroups *S*_1_ ⋃ *R*_1_:

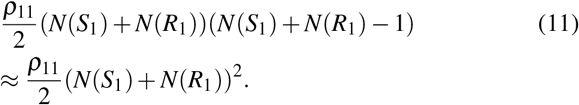

This number is going to appear in the dynamics of the state budget *W*:

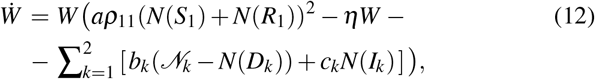

where *η* is the limiting factor, the term ∝ *a* stands for products and services created through the interactions in first group, i.e only the members of first group create public good.

*b_k_*, *k* = 1, 2 are standing for the spending. While the wealth growth is due to the interactions in the first group, the spending of wealth is due to both of the groups. We assume that the consumption rate is the same for both groups, *b*_1_ = *b*_2_.

*c_k_*, *k* = 1, 2 represent the additional cost of treatment of the infected people inside the hospitals, which will be elaborated later.

We assume that in the absence of infection, the wealth growth dynamics is governed by the logistic growth. The proposed growth model is known from population dynamics, and has also been used for modeling economic problems [24, 25, 26, 27, 28]. In the absence of any infection, the wealth growth dynamics have the following form

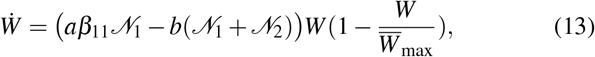

where we have used the following relation

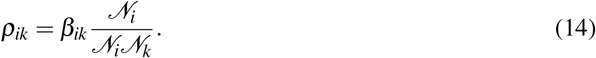

Eq.(13) is obtained from (12) by assuming that the limiting factor has the following form

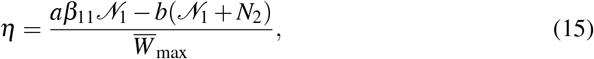

where 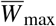 is the carrying capacity of wealth growth. Therefore, we assume that before the infection outbreak the wealth growth dynamics is given by logistic growth with constant reproduction rate 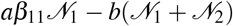 and capacity 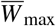. The quantities *a*, *b* and 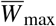 are free parameters and they have to be estimated from the economic situations of countries.

Hereafter, the frequencies of each subgroup will be denoted by the corresponding letter, i.e

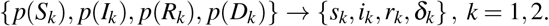

Let us denote the fraction of the first population over the total population by 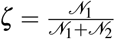.

In the case of a disease outbreak, the reproduction rate and the carrying capacity of wealth growth dynamics becomes time-dependent.

From (12) for the wealth growth dynamics we obtain the following equation

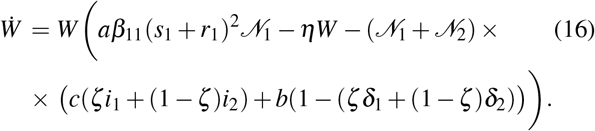

Note that here we assume that η is a constant, since factors that limit the economic growth are likely (at best) to stay constant after infection starts. Therefore, the change of the maximum possible value of wealth growth dynamics due to the disease outbreak may be found by considering the wealth growth dynamics in the long-run time scales.

In the long-run time scale, it is assumed that the infected fraction of the populations tend to zero, due to the recovery and/or death process *i_k_*_∞_ → 0*, k* = 1, 2. From (16) by assuming vanishing fractions of an infected population, we find the possible non-trivial rest point of the wealth growth dynamics, obviously, *W* = 0 is also a rest point for wealth growth dynamics. The non-zero rest point is found from (16) by nullifying the expression on the right hand-side and using the definition (15). We obtain:

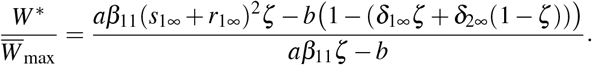

Here, 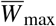 stands for the wealth growth capacity in the absence of infection outbreak. Note, that in the cases where *W*^*^ < 0, wealth tends to zero. Therefore, using the normalization of frequencies *s_k_*_∞_ + *r_k_*_∞_ + *δ_k_*_∞_ = 1*, k* = 1, 2, for the *W*\* > 0 case, this quantity reveals the maximum possible value of wealth growth dynamics *W*_max_ =*W*^*^. The fraction of the maximum values of wealth growth dynamics with and without disease outbreak, respectively *W*_max_ and 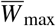, will be

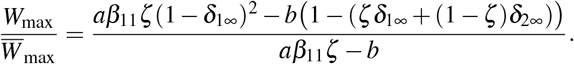

Thus, the disease outbreak may have different impacts on the wealth growth dynamics in the long-term. The fraction (17) can be both 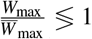 7 1. The expected impact depends on the structure of the total population, production and consumption rates, and the total size of the deaths in each population. For the COVID-19 disease outbreak, the fraction (17) is less than one.

### Feedback arising from economic growth and disease dynamics

Above we have described the impact of the infection transmission on the wealth growth dynamics.We assume that the wealth growth dynamics itself affects the infection dynamics due to the wealth dependency of death rates. The wealth dependency of death rates means that in the absence of possible resources, the treatment quality in the health care system will suffer. For the recovery rate *λ_k_* = *w*(*R_k_*|*I_k_*) *k* = 1, 2, we assume that it remains constant, i.e after the constant recovery period, the infected people either die or recover. We assume that the death rates *μ_k_* = *w*(*D_k_*|*I_k_*) *k* = 1, 2 are negatively correlated with the wealth growth inside the hospitals.

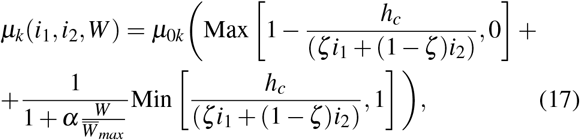

where *μ*_0_*_k_, k* = 1, 2 represent the natural death rates for each group outside the hospitals, and a represents the economic state of the population. Since we are going to describe the dynamics of the dimensionless quantity 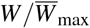, the difference between rich and poor countries will be lost.

In (17), the health system capacity is denoted by *h_c_*. Due to the health system capacity, only a certain portion of the infected population from each group will be treated in the hospitals. Indeed, when the total infected population does not exceed the health system capacity level *hc*, then the deaths in total population per-unit of time will be given by the following relation:

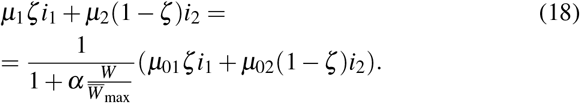

Otherwise, when the total fraction of infected population exceeds the health system capacity level, the latter quantity will be

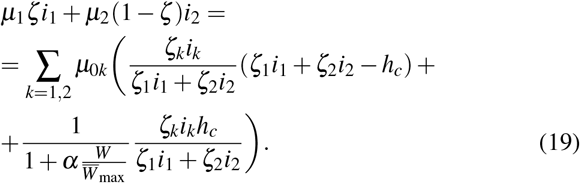

Here, *ζ*_1_ ≡ *ζ* and *ζ*_2_ ≡ 1 − *ζ*. Similar relations hold for the total deaths per unit of time.

Treatment cost in the hospitals is governed by the rate *c* in (12). Here again we assume that outside the hospitals the treatment cost is nullified. This means that people who are infected but cannot be treated in the hospitals (due to the capacity), will not “produce” additional costs. Hence, we have:

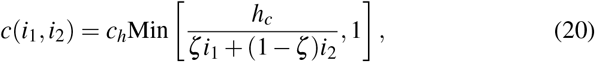

where *c_h_* is the rate of the cost of the treatment in hospitals.

### Calibration of parameters: Economic part

In order to determine relevant values for the parameters involving the economic part, we consider the wealth growth dynamics as a budget generating process in countries. Budget generation is based on the revenues and expenditures of a government. Though revenues are not solely based on the interaction of economic agents, it is reasonable to use the budget to obtain reliable estimates for parameters *a*, *b* and 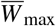.

As an example, we choose the budget dynamics of Germany. In 2017 the budget balance (difference of revenues and expenditures) of Germany was equal to 1.25%GDP_2017_ of that year [29]. The revenues Re_2017_ = 45%GDP_2017_ [30]. We use the real GDP of Germany in constant 2010 US$. According to theWorld Bank data, GDP_2017_ = 3.878 ×10^12^ US$ [31]. In 2018, the above mentioned quantities are 1.75% *GDP*_2018_, *GDP*_2018_ =3.937 × 10^12^ US$, *Re*_2018_ = 45.5%*GDP*_2018_. The relevant value for parameter *a* is obtained by assuming that revenue generation is given by the logistic growth:

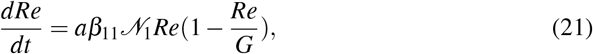

where *G* is the carrying capacity value and reproduction coefficient is 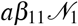. We assume that the revenue *Re*_2018_ is close to the carrying capacity *G* = 45.7%*GDP*_2018_ ≈ *Re*_2018_. This choice assumes that the revenues are collected as much as possible. From (21), it follows that the choice of time units is governed by the constant *a*, i.e a change in the unit of times will cause *a* to change. Therefore, we choose the day as the unit of time, since the parameters of the SIR model are defined in terms of days. The solution of (21) has the following form:

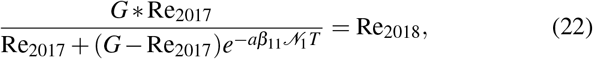

where *T* = 365 days. Total population of Germany in 2018 is equal to 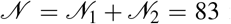 million people according to the World Bank data [32]. The age distribution was 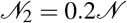, where 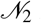 is the population size of those who are 65 and above. The contact number *β*_11_ = 10 (this choice will be explained below alongside with the other contact numbers *β*_12_ and *β*_22_). Hence, from (22), we estimate that the value of 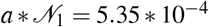. For the consumption rate, we introduce an equation similar to (22). We assume that the carrying capacity for expenditures is the same as for revenues. For expenditures (going through the reasoning above) we estimate the value of 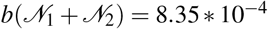. Thus, we obtain 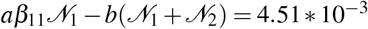 as the overall budget reproduction coefficient.

Here, we use the above estimation of budget parameters for (13). Note, that (13) does not govern the real budget dynamics now, since the real budget dynamics will be given by the difference of two logistic equations–revenues and expenditures. However, we use the parameters for (13) and try to find the carrying capacity level in order to provide the correct values for budget balance in 2017 and 2018. Indeed, using (13), we obtain an equation for 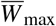

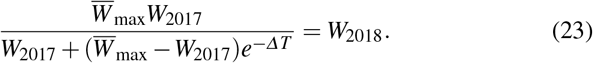

We have taken into account the fact that *W*_2017_ is defined through *GDP*_2017_. Solving the last equation, we find the value of the capacity of wealth growth dynamics 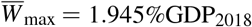 *GDP*_2018_.

### Calibration of parameters: Infection and health system part

In the classical SIR models, the disease outbreak dynamics is described by the reproduction number of disease R_0_ [14, 12, 13]. This quantity describes the number of secondary infectious cases caused by an infected individual when whole population is in suspected subgroups. The reproduction number varies during the spread of disease outbreak, since more people will be infected and hence less suspected population will remain. For the present model, the reproduction number can be found by expressing the equations of infected populations *i*_1_, *i*_2_ in matrix form [33, 34].

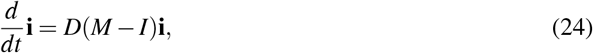

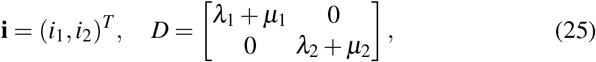

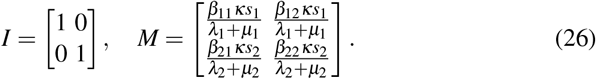

The reproduction number *R*_0_ defined as the maximum eigenvalue of matrix *M*, when *s*_1_*, s*_2_ ≈ 1.

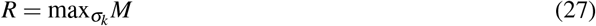

where *σ_k_*, *k* = 1, 2 are the eigenvalues of matrix *M*.

During the infection spread, the reproduction number *R* is decreasing. Let us now introduce the values of parameters involved in the matrix *M* describing Covid-19 disease outbreak. These parameters may vary due to further insights about the outbreak as time progresses. However, the scope of the present paper is to compare the different response strategies in relation to the given infection outbreak.

The recovery rates are assumed to be *λ*_1_ = *λ*_2_ = 1/18, i.e a period of 18 days is required for the transition from infected to recovered subgroup [1, 17, 35, 20]. During the recovery period people may die, which is described by the death rates. For the death rates outside the hospitals, we assume *μ*_01_ = 2 * 0.0025 * 1/*λ*_1_ and *μ*_02_ = 2 * 0.06 * 1/*λ*_2_. In the current literature, the death rates are assumed to be 0.0025 * 1/*λ*_1_ and 0.06 * 1/*λ*_2_ respectively for the first and second groups [17, 35]. These rates describe the deaths in the hospitals, i.e when infected patients receive medical treatment. That is why we assume that outside the hospitals, these parameters will increase. For illustration purpose, we assume that these parameters will increase by at least twice.

For the contact numbers, we use {*β*_11_, *β*_12_, *β*_22_} = {10, 6, 8}. According to the research undertaken to investigate the daily contacts (either skin-to-skin contact or two-side conversations) in different European countries [36], the average number of contacts in Germany was 7.95, so we assumed an average of 8. Meanwhile, the contacts follow an age-dependent pattern, i.e people within the same age group contact one another more often than people from outside their age ground.

It is worth noting that in the classical SIR-model, the fraction of deaths is not explicitly included. Hence, the reproduction number may vary depending on the way the deaths are accounted for in the model. For the Covid-19, the reproduction number *R*_0_ is assumed to be *R*_0_ ≈ 2.4 [1]. However, the deaths caused by infection have not been explicitly included in this estimation. We choose *κ* = 0.009 as to match this value without deaths, viz. *R*_0_ ≈ 2.44. In the presence of deaths the reproduction number is *R*_0_ ≈ 2.38.

For the health system capacity level, we assume *h_c_* = 0.01, i.e the health system is capable to provide treatment to the 1% of the population. Indeed, this value seems much greater than the exact capacities of health system in most countries. Therefore, we have also taken into account the fact that not all of the infected people need hospitalization [1]. Treatment cost has a temporal effect on the wealth dynamics, viz. changing the treatment cost varies the depth of economic recession. We choose a representative value *c* = 10*b*. All the parameters and their values are presented in the table below.

|  |  |  |  |
| --- | --- | --- | --- |
| $\mathcal{N}$ | $83 * 10^6$ | $\lambda_1, \lambda_2$ | $1/18$ |
| $\mathcal{N}_1$ | $0.8 * \mathcal{N}$ | $\mu_{01}$ | $2 * 0.0025 * 1/18$ |
| $\mathcal{N}_2$ | $0.2 * \mathcal{N}$ | $\mu_{02}$ | $2 * 0.06 * 1/18$ |
| $GDP_{2018}$ | $3.937 * 10^{12}$ | $\kappa$ | $0.009$ |
| $W_0$ | $1.75\% GDP_{2018}$ | $h_c$ | $0.01$ |
| $\bar{W}_{max}$ | $1.945\% GDP_{2018}$ | $c$ | $10b$ |
| $a * \mathcal{N}_1$ | $5.35 * 10^{-4}$ | $\beta_{11}$ | $10$ |
| $b(\mathcal{N}_1 + \mathcal{N}_2)$ | $8.35 * 10^{-4}$ | $\beta_{12}, \beta_{21}$ | $6$ |
| | | $\beta_{22}$ | $8$ |

## Results

### Mitigation scenarios

We now discuss three different responses to the disease outbreak. We assume that neither a vaccine nor effective antiviral drugs become available. We will also assume that the contact numbers can be reduced by discrete values. We assume that the response to the disease started at the point in time when the fraction of the infected population exceeded some threshold value 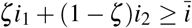. Response strategies are defined as follows:

1. Total lockdown: Here we assume that all the contact numbers are reduced by a discrete value: specifically, they reduce by five times.
2. Partial lockdown: Here we assume that the contact numbers *β*_12_ and *β*_22_ will be reduced, while *β*_11_ remains constant.
3. Towards herd immunity: In this scenario, again, *β*_11_ remains constant. However, *β*_12_ and *β*_22_ are chosen such that enough of the population has been infected during the mitigation, and that the infection will not grow again when measures are relaxed.

The results arising from the above strategies will be compared against the “do-nothing” scenario, i.e no mitigation measures are being imposed.

### Total lockdown strategy

We now discuss the total lockdown scenario as a mitigation measure, where the goal is to reduce the reproduction number of the disease (27) to below one. In the context of our model, the end-point of the total lockdown scenario is not clear in the absence of a vaccine. As an end-point of the mitigation measures, we choose the moment in time when the total fraction of the infected population is reduced to the initial value of the infection (starting point of disease dynamics). We assume that all contact numbers are reduced by five times: 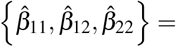 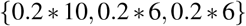.

The wealth growth dynamics, infection spread and the fraction of deaths are represented in Fig.1. In the left panel (Fig.1a), the wealth growth dynamics is represented under the mitigation measures (full line), “do nothing” scenario (dashed line) and in the absence of disease outbreak (dotted line). Under the “do nothing” scenario, the wealth is decreasing due to the infection of the first population and due to the treatment cost of the infected population, as it follows from (13). Furthermore, the maximum possible value of wealth defined by the relation (17) is decreased 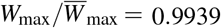.

**Fig. 1:**
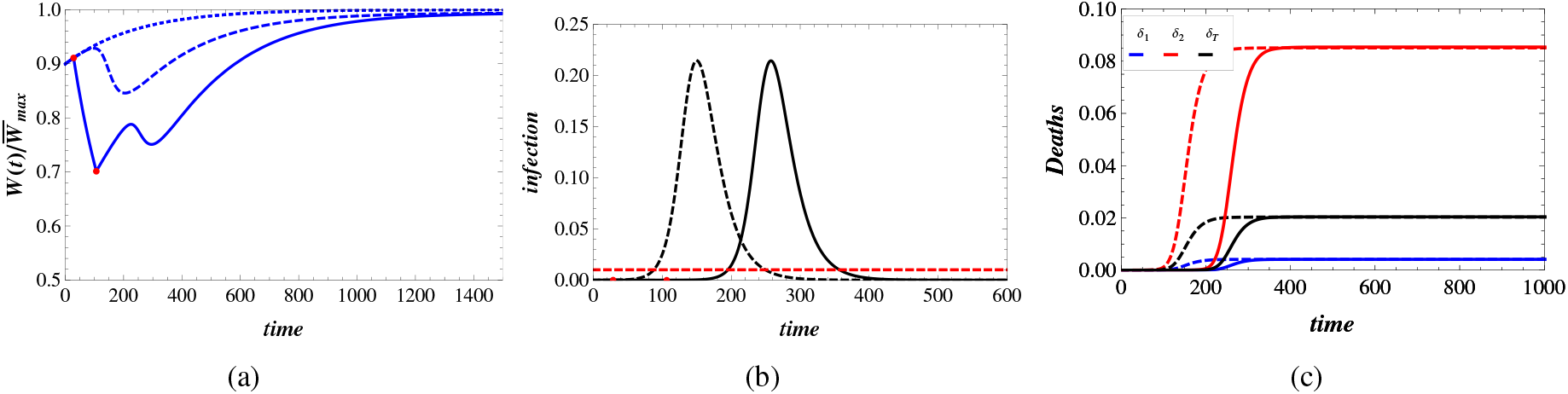
Total lockdown and “do-nothing” scenarios. The contact numbers during the total lockdown strategy are 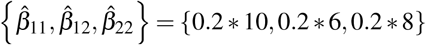. Dynamics of the fraction of wealth and its capacity level Fig.1a, infection spread Fig.1b and the dynamics of deaths Fig.1c. In the left panel (Fig.1a), the dashed (dotted) blue line represents the wealth dynamics in the “do-nothing” (in the absence of infection) scenario. The full blue line represents wealth growth dynamics under total lockdown and after that. Red points represent the start and the end of lockdown. The full (dashed) black line on the middle panel (Fig.1b) represents infection spread under total lockdown (”do nothing”) scenario and after that. Red dashed line represents health system capacity level. In the right panel (Fig.1c), the full blue, red and black lines represent the fraction of deaths in the respective first, second, and in total population (*δ_T_* ≡ *ζδ*_1_ +(1 − *ζ*)*δ*_2_). Dashed lines illustrate the behavior of these quantities in the “do-nothing scenario”. Initial values for the spread dynamics are *i*_1_ = *i*_2_ = 10^−5^. The mitigation measures are imposed at the value 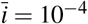.

The first red point on the full line in (Fig.1a) illustrates the start point of the lockdown measures—when the total infection 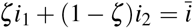. The second red point in (1a) shows the moment of lifting the measures—when the total infection becomes equal to the starting value of infection *ζi*_1_(0)+(1 − *ζ*)*i*_2_(0)=10^−5^. The wealth growth comes down drastically during the lockdown period, since the growth is reliant on the contact number *β*_11_ (which is reduced by five times during the lockdown).

During the lockdown period, the fraction of the infected population is decreasing. After the lifting of the lockdown measures, the disease outbreak does not go extinct (there is no such a mechanism as part of our model), and it starts to spread again after some time. During this interval, wealth will start growing. Then, it starts decreasing again, however, this time due to the infection spread. In this case, the minimum value of wealth dynamics is smaller than in the “do nothing” scenario. This means that the death rates become greater (i.e the quality of treatment decreased) in the hospitals compared to the worse situation in “do nothing” scenario. The fraction of deaths is illustrated in Fig.1c. As can be seen, the fraction of deaths is increased, since the quality of treatment is worsened. In our context, the discrepancy is negligible. Thus, the resource depletion can cause more deaths, and this phenomenon observed for the general Susceptible-Infected-Susceptible (SIS) model that has been discussed in [37]. The fraction of deaths in the two populations *δ*_2∞_ = *δ*_1∞_ is approximately the same for both scenarios, but is larger in the “do nothing” scenario. Due to this and the greater fraction of deaths of the first population in the total lockdown strategy, the maximum value of wealth in the total lockdown strategy is smaller than in the “do nothing” scenario 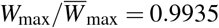.

### Partial lockdown

The consequences of the total lockdown strategy can be more unfavourable for the population than in the “do-nothing” scenario, as we have seen above. The major reason for this phenomenon is the depletion of the resources during the total lockdown. In the partial lockdown strategy, the goals differ from the above-discussed case in two aspects. Here, in the partial lockdown, the public health goal is to preserve the total fraction of the infected population to be as low as possible. Meanwhile, we have to prevent the large recession in wealth dynamics, which occurs in total lockdown strategy. As can be seen from (12), the wealth growing process is based on the contacts in the first group *β*_11_. Thus, for keeping up the wealth situation we assume that the mitigation measures do not affect *β*_11_. We choose the value of contact numbers as follows: 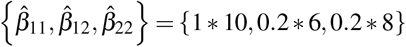. In the partial lockdown case, the starting and ending times of mitigation are defined in the same way as in the total lockdown scenario.

The behavior of the wealth dynamics, infection spread and the fraction of deaths is presented in Fig.2. The first period of infection does not induce as great a damage to the economy as in the “do nothing” scenario. Meanwhile, at the start of the second wave—which occurs because there is no vaccine and not enough people have been infected in the mitigation phase—the wealth of the population is in a better situation than at the start of the infection.

**Fig. 2:**
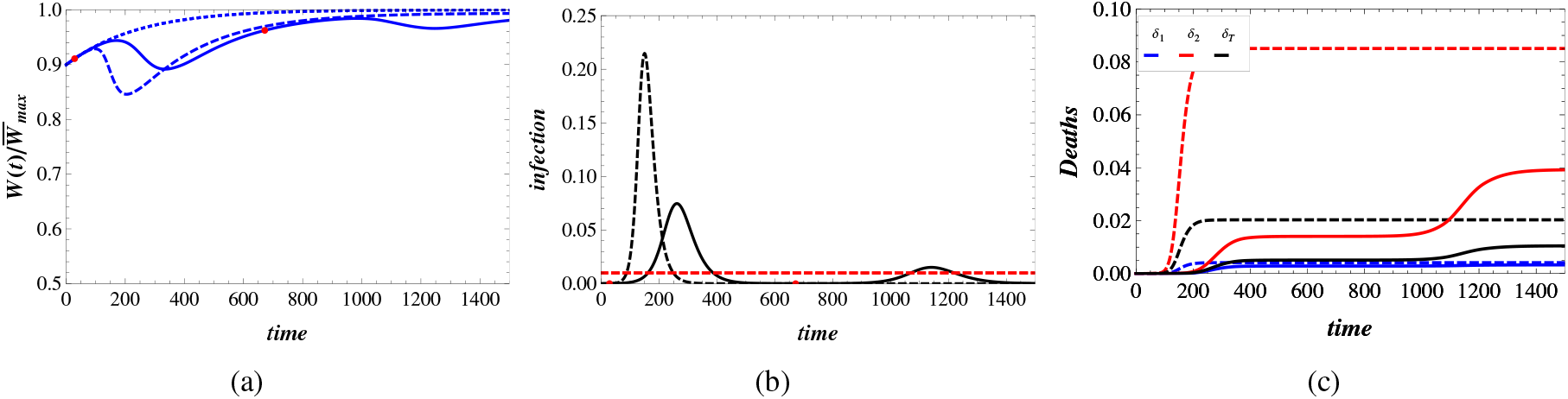
Partial lockdown 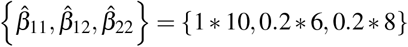 and “do-nothing” scenarios. Description of the figures are the same as in Fig.1.

This means that the treatment quality is at least as good as at the start of the epidemic. Furthermore, during the second wave of the infection spreading, wealth dynamics do not decrease as much as in the “do nothing” scenario. Due to the fact that at the start of the second wave, part of the population has been infected already and the quality of treatment in the hospitals become better than at the start of the infection. The behavior of the fraction of the total infected population under partial dock-down and “do nothing” scenarios are presented in Fig.2b. It is seen that more people are treated inside the hospitals in partial lockdown strategy than in “do nothing scenario”. Due to these situations, the fraction of deaths is drastically reduced under the partial lockdown strategy as shown in Fig.2c. The total number of deaths is almost twice as small under the partial mitigation measures than in the “do nothing” scenario.

The maximum possible value of wealth under the partial lockdown is approximately the same as in the absence of disease 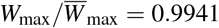. Thus, for both public health and economy parts, the partial lockdown strategy minimizes the losses.

It is necessary to mention, that we are discussing the one-round lockdown strategies, i.e the mitigation measures are imposed at once. For example, under the partial lockdown strategy, the second wave of the disease can be suppressed by increasing health system capacity. However, these questions are beyond the scope of the current paper.

### Towards herd immunity

The next strategy of response to the disease outbreak is a mitigation of the spread dynamics to ensure the occurrence of herd immunity in the population. In the classical SIR model, the occurrence of herd immunity (here it has to be mentioned that occurrence of herd immunity is based on, and a consequence of the epidemic model, in our case it is the SIR model) is associated with the peak of the infection curve, i.e the point after which infection goes to extinction. Thus, the “do nothing” scenario is also providing the herd immunity in the population. However, the “do-nothing” scenario is not the optimal strategy for achieving our goal for at least two reasons: health system capacity and wealth dynamics.

The reproduction number (27) has to be small enough in order to slow down the spread of the infection. In this way, more infected people will be treated in hospitals. Therefore, it has to be large enough so as to ensure the disease transmission is in the population. The difference between the present strategy and partial lockdown is the absence of the second wave of the infection after the relaxation of the mitigation measures. As in the previous cases, we assume here that the start of the mitigation measures is at the instance of time when the fraction of the total infected population exceeds the threshold value 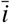. The endpoint of the mitigation measures *t*^*^ is defined from the following consideration: after relaxation of the mitigation measures—when contact numbers become the same as before the start of the mitigation process—the infection spread has to decline. Thus, the mitigation measures are lifted whenever the following condition is satisfied:

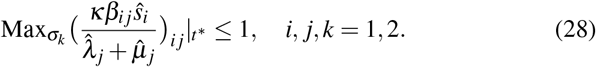

Here *ζ_k_* are the eigenvalues. In (28), the contact numbers are given by those in the pre-mitigation period, while the dynamics of infection is controlled by the contact numbers in the mitigation period 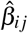. The contact numbers in the mitigation period are 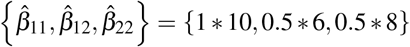. We assume that the contact numbers in the first group remain the same as before the mitigation period.

Infection spread under the “towards herd immunity” and “do nothing” strategies is illustrated in the Fig.3b. As can be seen from Fig.3b, the time at which the fraction of infected people exceeds the health system capacity level, is greater than in the “do nothing” scenario. Indeed, when the fraction of infected population exceeds health system capacity, the reproduction number of disease at that moment of time is greater in the “do nothing” scenario. This is because at that moment of time, fewer people remain suspected in the present strategy than in the “do nothing” scenario. Meanwhile, more people have been treated in hospitals in the mitigation regime.

**Fig. 3:**
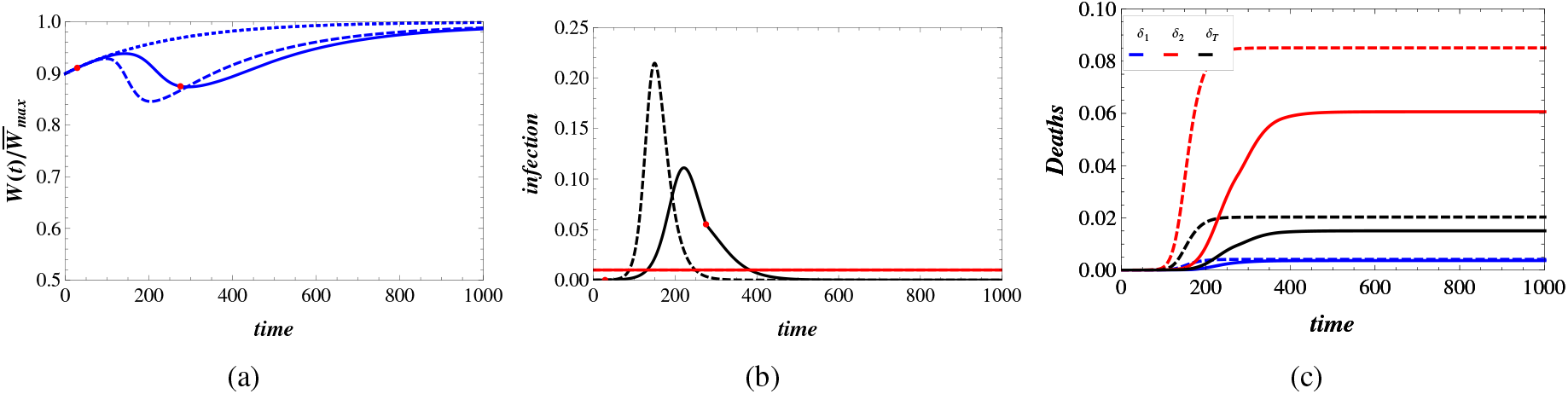
Towards herd immunity by mitigation measures and without them, i.e “do nothing” strategy. 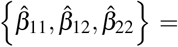 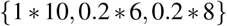. Wealth growth Fig.3a, infection spread Fig.3b and deaths Fig.3c. Description of the figure is the same as in Fig.1

The dynamics of wealth and the fraction of deaths are represented in Fig.3a and Fig.3c respectively. The wealth grows in the initial phase of the epidemic is seen in Fig.3a. The total fraction of deaths in the present scenario is slightly greater than the total deaths under the partial lockdown strategy. The maximum possible value of wealth under the “towards herd immunity” strategy is approximately the same as in the partial lockdown strategy 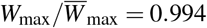. Comparing the results from partial lockdown and towards herd immunity strategies, it becomes clear that the deaths caused by infection spread is lesser in the partial lockdown strategy. Meanwhile, we assume that the vaccination is not yet available. If at some point in time during the mitigation measures the vaccine is being made available, then the partial lockdown strategy is more favorable than the other strategies since more people will be vaccinated. The point is that in the partial lockdown strategy, the second wave of infection will occur, in contrast to the total lockdown and towards herd immunity strategies. Thus, partial lockdown strategy divides the expected wave of disease outbreak in two parts and by this way ensures that more people will be treated inside the hospitals with better treatment quality. Therefore, it is necessary to mention that the duration (right and left red points on the figures) of partial lockdown strategy is greater than for the other strategies.

## Discussion

We will now discuss the impact of the different strategies in response to the disease outbreak with regards to the wealth growth and disease transmission dynamics. We assume that neither a vaccine nor effective antiviral drugs become available. To this end, we discuss disease outbreak transmission in an age-structured population: young and old groups. The latter group is considered more vulnerable to the infection: the death rate is higher for this group. We have chosen the parameters based on real-world data related to the COVID-19 pandemic in this paper. We have discussed three different response strategies to the pandemic in the absence of vaccine and effective antiviral drugs: total lockdown, partial lockdown, and towards herd immunity. We assume that the mitigation measures are lifted whenever the final goal of the strategies is obtained. We compare the deaths in the population and the economic impact of the mitigation strategies with a “do-nothing” strategy, namely, when there are no mitigation measures at all.

The total lockdown strategy tries to decrease the reproduction number of disease below one, that is, to suppress the further transmission of disease inside the community. The contact numbers in each group and the contact numbers of intergroup interactions are reduced by the same value during the total lockdown period. The number of infected people decreases during the mitigation period, so the overall treatment cost decreases. The reason behind the economy worsening is the reduced intragroup interactions in the first group.

The endpoint of the total lockdown strategy in the absence of a vaccine is supposed to be the instance when the fraction of infected people declines to below the threshold value. After relaxing the mitigation measures, the disease outbreak continues to spread in the society. However, in the context of our model, the consequences can be more severe than in the “do-nothing” scenario. Under the considered parameters range, the difference between the total deaths at the end of COVID-19 pandemic is small when these two strategies are implemented; In the long run, the maximum possible value of wealth is changed because of the deaths. Here, again, the total lockdown strategy is not favored in the absence of a vaccine.

The next strategy of response to the disease outbreak is a partial lockdown. The goals of the strategy differ from total lockdown in two ways: on the one hand to slow down the disease transmission in the society so that infected patients can obtain the required treatment in the hospitals as much as possible, and on the other hand to keep the wealth growth dynamics to an accepted level. For the latter purpose, the intragroup contact number of the first group members remains constant. The other two contact numbers: intergroup contacts and intragroup contacts in the second group, will decrease by the same amount as in the total lockdown strategy.

Under our considered parameters, the second wave of disease outbreak will occur after lifting the mitigation measures. However, at this time, the peak of the infection is smaller than in the first phase of infection. The total deaths in the second group population are approximately twice smaller than in the “do-nothing” scenario. This is because of three reasons: firstly, people from the vulnerable group are infected less than in the “do-nothing” scenario, so the fraction of deaths decreases. Secondly, infected people receive relatively good treatment inside the hospitals. Lastly, resources are increasing before the second wave, hence the quality of the treatment inside the hospitals can be increased. The deaths of the first group are approximately the same for both the partial lockdown and “do nothing” scenarios since the intragroup interactions in the first group remain at the same level. Thus, the fraction of total deaths in the whole population Population is approximately twice smaller in the partial lockdown scenario compared with the “do nothing” scenario. In this case, the decline in wealth dynamics is caused by the infection spread only: infected people do not contribute to the wealth growth process, in contrast to the previous scenario where the resource depletion occurs before the peak of the epidemic. Therefore, the economic decline in the partial lockdown strategy is not as severe as in the “do nothing” scenario due to the slowdown in the disease transmission. The long-term impact of the infection outbreak is also milder for the partial lockdown strategy.

The last scenario for the response of the infection outbreak (in the absence of the vaccine) is a strategy towards herd immunity. It should be noted that the occurrence of herd immunity is based on the dynamical properties of SIR model, as we have assumed that the recovered people will not be infected again. Here we emphasize that the “do nothing” strategy also guarantees the occurrence of herd immunity in the population: a turnover point in the infection dynamics after which infection declines and goes to extinction. However, this is not the favoured way to build herd immunity under the health system capacity levels and the economic consequences of infection transmission. As such, we discuss the following strategy for building herd immunity in the population: the intragroup interaction between the members of the first group remains at the same level, while the other two contact numbers decrease by an amount such that the infection has to continue spreading in the population, but in contrast to the partial lockdown case, a second wave of the infection may not appear in the future. The endpoint of the mitigation measures is defined by the threshold value of the reproduction number: when relaxing the mitigation measures, this will not cause a growth in the fraction of the infected population.

Due to the decrease in contact numbers, the deaths caused by infection is smaller in this scenario than in the “do nothing” case. The reason behind the decline in deaths is the same as in the partial lockdown case, but the total case of deaths in the partial lockdown is smaller than in the herd immunity scenario. As in the partial lockdown scenario, the economic situation during the outbreak in the herd immunity scenario is better than in the “do nothing” scenario in both short and long-term.

## Conclusion

In summary, we have taken a ‘realpolitik’ approach by exploring mortality rates and economic considerations in the same model together as it is done in reality. In our theoretical model, we have assumed that neither a vaccine nor effective antiviral drugs become available. Based on the context of our study, it is clear that the partial lockdown strategy is a more preferable choice compared to the other two strategies, in terms of public health and economic considerations. Finally, a word of caution: COVID-19 is a complex of medical conditions, and not a cause. The ultimate causative agent is not a virus in isolation, but a virus in complex with particular social factors – the Four Horsemen: a) Overpopulation, b) Globalization, c) Hyperconnectivity, and d) Extreme centralization and increasing fragility of supply chains [38]. The fact is that we are in an unsustainable geosocial situation. If these conditions are not resolved, we should consider recurrent catastrophe as an inevitable emergent phenomenon of the dynamics that emerge from such complex systems.

## Data Availability

All data are included in the manuscript.

## References

1. N. Ferguson, D. Laydon, G. Nedjati Gilani, N. Imai, K. Ainslie, M. Baguelin, S. Bhatia, A. Boonyasiri, Z. Cucunuba Perez, G. Cuomo-Dannenburg, et al., (2020)

2. A. De Visscher, Nonlinear Dynamics (2020)

3. P.G. Walker, C. Whittaker, O.J. Watson, M. Baguelin, P. Winskill, A. Hamlet, B.A. Djafaara, Z. Cucunubá, D.O. Mesa, W. Green, et al., Science (2020)

4. A.R. Tuite, D.N. Fisman, A.L. Greer, CMAJ 192(19), E497 (2020)

5. S.H. Ebrahim, Q.A. Ahmed, E. Gozzer, P. Schlagenhauf, Z.A. Memish. Covid-19 and community mitigation strategies in a pandemic (2020)

6. R.M. Anderson, H. Heesterbeek, D. Klinkenberg, T.D. Hollingsworth, The Lancet 395(10228), 931 (2020)

7. R. Singh, R. Adhikari, arXiv preprint arXiv:2003.12055 (2020)

8. C. Kwuimy, F. Nazari, X. Jiao, P. Rohani, C. Nataraj, Nonlinear Dynamics pp. 1–15 (2020)

9. S. He, Y. Peng, K. Sun, Nonlinear Dynamics pp. 1–14 (2020)

10. X. Wang, S. Wang, Y. Lan, X. Tao, J. Xiao, Nonlinear Dynamics pp. 1–10 (2020)

11. W.O. Kermack, A.G. McKendrick, Proceedings of the royal society of london. Series A, Containing papers of a mathematical and physical character 115(772), 700 (1927)

12. M.J. Keeling, P. Rohani, Modeling infectious diseases in humans and animals (Princeton University Press, 2011)

13. M. Martcheva, An introduction to mathematical epidemiology, vol. 61 (Springer, 2015)

14. H. Hethcote, Lecture Notes Series, Institute for Mathematical Sciences, National University of Singapore 16, 1 (2008)

15. S.R. Baker, N. Bloom, S.J. Davis, S.J. Terry, Covid-induced economic uncertainty. Tech. rep., National Bureau of Economic Research (2020)

16. A. Brodeur, D.M. Gray, A. Islam, S. Bhuiyan, (2020)

17. D. Acemoglu, V. Chernozhukov, I. Werning, M.D. Whinston, A multi-risk sir model with optimally targeted lockdown. Tech. rep., National Bureau of Economic Research (2020)

18. M.S. Eichenbaum, S. Rebelo, M. Trabandt, The macroeconomics of epidemics. Tech. rep., National Bureau of Economic Research (2020)

19. F.E. Alvarez, D. Argente, F. Lippi, A simple planning problem for covid-19 lockdown. Tech. rep., National Bureau of Economic Research (2020)

20. A. Atkeson, What will be the economic impact of covid-19 in the us? rough estimates of disease scenarios. Tech. rep., National Bureau of Economic Research (2020)

21. P. Carlsson-Szlezak, M. Reeves, P. Swartz, Harvard Business Review 3 (2020)

22. R. Chang, A. Velasco, Economic policy incentives to preserve lives and livelihoods. Tech. rep., National Bureau of Economic Research (2020)

23. P. Carlsson-Szlezak, M. Reeves, P. Swartz, Harvard Business Review (2020)

24. S. Girdzijauskas, D. Štreimikiene, Journal of Business Economics and Management 10(1), 45 (2009)

25. W. Kwasnicki, Technological Forecasting and Social Change 80(1), 50 (2013)

26. G.P. Boretos, Technological Forecasting and Social Change 76(3), 316 (2009)

27. T. Modis, Technological Forecasting and Social Change 80(8), 1557 (2013)

28. S.E. Puliafito, J.L. Puliafito, M.C. Grand, Ecological Economics 65(3), 602 (2008)

29. Budget balance to gdp. https://www.statista.com/statistics/624187/germany-budget-balance-in-relation-to-gdp/

30. 2018 draft budgetary plan germany. https://ec.europa.eu/info/sites/info/files/2018_dbp_de.pdf

31. Gdp (constant 2010 us $). https://data.worldbank.org/indicator/NY.GDP.MKTP.KD?locations=DE

32. Population. https://data.worldbank.org/indicator/SP.POP.TOTL?end=2018&locations=EU-DE&start=2009

33. H. Guo, M.Y. Li, Z. Shuai, Canadian applied mathematics quarterly 14(3), 259 (2006)

34. P. Magal, O. Seydi, G. Webb, SIAM Journal on Applied Mathematics 76(5), 2042 (2016)

35. A. Glover, J. Heathcote, D. Krueger, J.V. Ríos-Rull, Health versus wealth: On the distributional effects of controlling a pandemic. Tech. rep., National Bureau of Economic Research (2020)

36. J. Mossong, N. Hens, M. Jit, P. Beutels, K. Auranen, R. Mikolajczyk, M. Massari, S. Salmaso, G.S. Tomba, J. Wallinga, et al., PLoS Med 5(3), e74 (2008)

37. L. Böttcher, O. Woolley-Meza, N.A. Araújo, H.J. Herrmann, D. Helbing, Scientific reports 5(1), 1 (2015)

38. K.H. Cheong, M.C. Jones, BioEssays 42, 2000063 (2020)

